# Assessing the value of linking public health microbiology data to the UK Biobank

**DOI:** 10.1101/2025.07.01.25330627

**Authors:** Shang-Kuan Lin, Jacob Armstrong, Amanda Y. Chong, Guillaume Butler-Laporte, Naomi E. Allen, Alexander J. Mentzer, David H. Wyllie, Daniel J. Wilson

## Abstract

Infection is important both as a cause of communicable diseases and as an exposure predisposing to non-communicable diseases. Investigating disease risk is a major research focus in large cohorts like UK Biobank. Linking cohorts to electronic health records, like the UK Health Security Agency’s Second Generation Surveillance System (SGSS), can enhance infection research. SGSS collects infection data from ∼200 microbiology laboratories across England, supporting surveillance, outbreak detection, and antimicrobial resistance monitoring.

We previously described algorithms linking SGSS to UK Biobank and demonstrated their utility during the COVID-19 pandemic. Here, we assess the value of SGSS for infection research by comparing it to Hospital Episode Statistics (HES), a centralized clinical dataset on hospital admissions already available in UK Biobank. Genome-wide association studies (GWAS) were used to evaluate the performance of SGSS microbiological diagnoses versus HES diagnostic codes for identifying infection outcomes.

SGSS contained substantially more infection records than HES by participant (82,888 vs 18,054), particularly for bacteria (excepting *Helicobacter pylori* and *Mycobacterium tuberculosis*). SGSS yielded more GWAS hits (31 vs 12) encompassing more pathogens (12 vs 8). Our findings demonstrate the high scientific added-value of SGSS above and beyond that of HES, supporting its integration in UK Biobank for future infection research.

## Introduction

Infection is an important cause of mortality and morbidity globally. Even in countries like the UK, where the burden of infectious diseases is relatively low compared to other parts of the world, pre-pandemic estimates indicate that infections accounted for 7% of all deaths, millions of lost workdays, and an economic impact of £30 billion per year^1–3^. Respiratory, bloodstream, and intestinal infections remain leading killers^4^, while sepsis, healthcare-acquired infections and antimicrobial resistance present ongoing challenges^5^. Infection poses a particular threat to infants and the elderly^6,7^.

Infection contributes to disease burden not only directly, through the spread of communicable disease, but indirectly, by predisposing to non-communicable diseases^8^. Infection with pathogens such as *Helicobacter pylori*, *Streptococcus pyogenes, Campylobacter jejuni*, Hepatitis C Virus and Human Papilloma Virus induce oncologic, inflammatory, auto-immune and neurological diseases including gastric^9^ and cervical cancer^13^ rheumatic heart disease^10^, Guillain-Barré syndrome^11^ and liver cirrhosis^12^. Infection is also implicated in conditions including type II diabetes^14^, chronic obstructive pulmonary disease^15^, and dementia^16^. Large longitudinal cohort studies can help untangle these often-complex relationships between infection and chronic disease, if high-quality data on infection and subsequent health outcomes are available.

UK Biobank (UKB) is a large prospective cohort study that collects comprehensive health, lifestyle, environmental and genetic data on 500,000 participants who were aged between 40-69 years at recruitment^17^. Participants lived near one of 22 recruitment centres between 2006 and 2010. UKB is among the world’s most important cohorts used by researchers internationally to investigate the determinants of health outcomes in mid-to-late life and their underlying biological mechanisms ^18–20^. All participants consented for their health to be followed long-term via linkage of their health-related records, including primary care, hospital, cancer and mortality records. A notable strength of UKB has been the continual enhancement of the data following recruitment, such as web-based questionnaires^21^, biomarker data (including biochemistry^22^, metabolomics^23^, proteomics^24^, telomere length^25^ and infectious disease markers^26,27^), and genotyping, whole exome^28^ and whole-genome^29^ sequencing of the whole cohort.

Currently, the identification of infection events in UKB relies primarily on linkage to hospital inpatient admissions data (which includes critical care)^30,31^. Diagnoses are professionally coded post-discharge using medical records, employing the International Classification of Diseases, Tenth Revision (ICD-10)^32^. Like any dataset repurposed for research, attention to artefacts caused by temporal or geographic idiosyncrasies is advised^33,34^. Commonly, hospital diagnoses of infection are based on clinical presentation. Microbiological investigation may determine the causative organism, but details are not necessarily recorded in hospital admission records and many diagnoses are recorded using ICD-10 non-specific codes like A41.0 “Septicaemia, unspecified”.

More specific diagnoses could be obtained in England – which covers 89% of the cohort – by linking to the United Kingdom Health Security Agency (UKHSA)’s Second Generation Surveillance System (SGSS). SGSS collects infection data from some 200 NHS laboratories serving general practices and hospitals across England to support surveillance, outbreak detection and antimicrobial resistance monitoring. SGSS comprises (i) the Communicable Disease Reports, which record mostly bacterial and viral pathogens in Schedule 2 of the Health Protection (Notification) Regulations 2010, and (ii) the Antimicrobial Resistance Reports (AMR), which record all bacteria with antimicrobial test results^35^. The bulk of NHS microbiology laboratories’ work involves culturing bacterial pathogens, and routinely testing them for antimicrobial resistance. Therefore data on bacterial pathogens described by the AMR table is most likely to improve infection diagnoses by specifying the causative organism and the specimen type (e.g. blood or urine).

Previously we described algorithm development for linking SGSS to UKB^7,36^. For January 2015 onwards, we linked an estimated 98% of infection events affecting UKB participants in England, ^7^. In 2020, laboratory-confirmed SARS-CoV-2 tests were linked to SGSS. Under a Control of Patient Information (COPI) notice issued by the UK government, SGSS-UKB linkage was deployed to allow data on SARS-CoV-2 testing to be made available to UK Biobank to enable researchers to study SARS-CoV-2 infection, with the first tranche of data released on 17^th^ April 2020^36^.

Laboratory-confirmed SARS-CoV-2 infection data, together with data on SARS-CoV-2 infection from hospital inpatient and death records enabled researchers to study the determinants of COVID-19 across the full spectrum of disease severity. More than 120 publications have investigated over 40 risk factors for COVID-19 outcomes in UKB participants (tabulated in ^37^).

Genome-wide association studies (GWAS) of COVID-19 outcomes in UKB participants contributed to meta-analyses that revealed over 50 regions of the human genome conferring susceptibility to SARS-CoV-2 infection, hospitalization, or critical COVID-19^38–41^. These studies underline the potential impact of linking SGSS to UKB to facilitate research into the genetic, lifestyle and environmental determinants of other infections.

In this study, we quantified the value of linking infection records from SGSS to UKB, beyond the information already available through Hospital Episode Statistics (HES), which are hospital inpatient records that cover England, for pathogens other than SARS-CoV-2. We evaluated the proposition that SGSS data could enhance our understanding of infection by providing more numerous and more specific case definitions of infection events than HES data. To test the added scientific value of SGSS data for case ascertainment, we focused on genetic risk factors for various infections, allowing us to (i) avoid concerns over mediation and, (ii) mitigate against confounding. Mediation – in which a risk factor influences the outcome indirectly through other risk factors – is less of a concern in GWAS because genetic variants are determined at conception and not influenced by downstream traits. Confounding – where an external factor, like ancestry, influences both the variant and the outcome – can still arise, but mature methods that adjust for population structure are effective in controlling it. For each pathogen, we summarized the numbers of infection events and genome-wide significant associations derived from SGSS vs HES. By analysing the broad picture and identifying important exceptions, we better characterize the utility of SGSS for studying infection in UKB.

## Results

### Identifying infection events in SGSS vs HES

We identified UKB-linked infection records in SGSS using the AMR table, which accounts for the bulk of records in SGSS, and mainly focuses on bacteria diagnosed by culture-based methods. For each record, we had information on the participant, organism identified, specimen date and the specimen type. We identified infection records in HES using the HESIN and HESIN_DIAG tables. Each record represented one episode, defined as a continuous period of hospital care under one consultant. We retained all episodes associated with infection-related primary or secondary diagnosis codes. SGSS records covered only England, with 99.9% of records from 3^rd^ December 2008 until 12^th^ November 2021, whereas HES records covered England, Wales and Scotland, with 99.9% of records from 8^th^ April 1997 until 7^th^ May 2021.

Figure 1 shows there were 349,612 versus 172,154 infection records derived from SGSS versus HES, a ratio of 1.9-fold. These numbers are restricted to participants resident in England with complete follow-up. For both SGSS and HES, a single infection event can give rise to multiple infection records. Therefore we deduplicated infection records by UKB participant, implemented by ignoring all but the first report per participant of any microbial taxon. De-duplicating this way, 114,292 versus 63,657 UKB participants had at least one infection diagnosis derived from SGSS versus HES, a ratio of 1.6-fold. Of those, 32,354 participants received the same infection diagnosis by both SGSS and HES.

**Figure 1.**
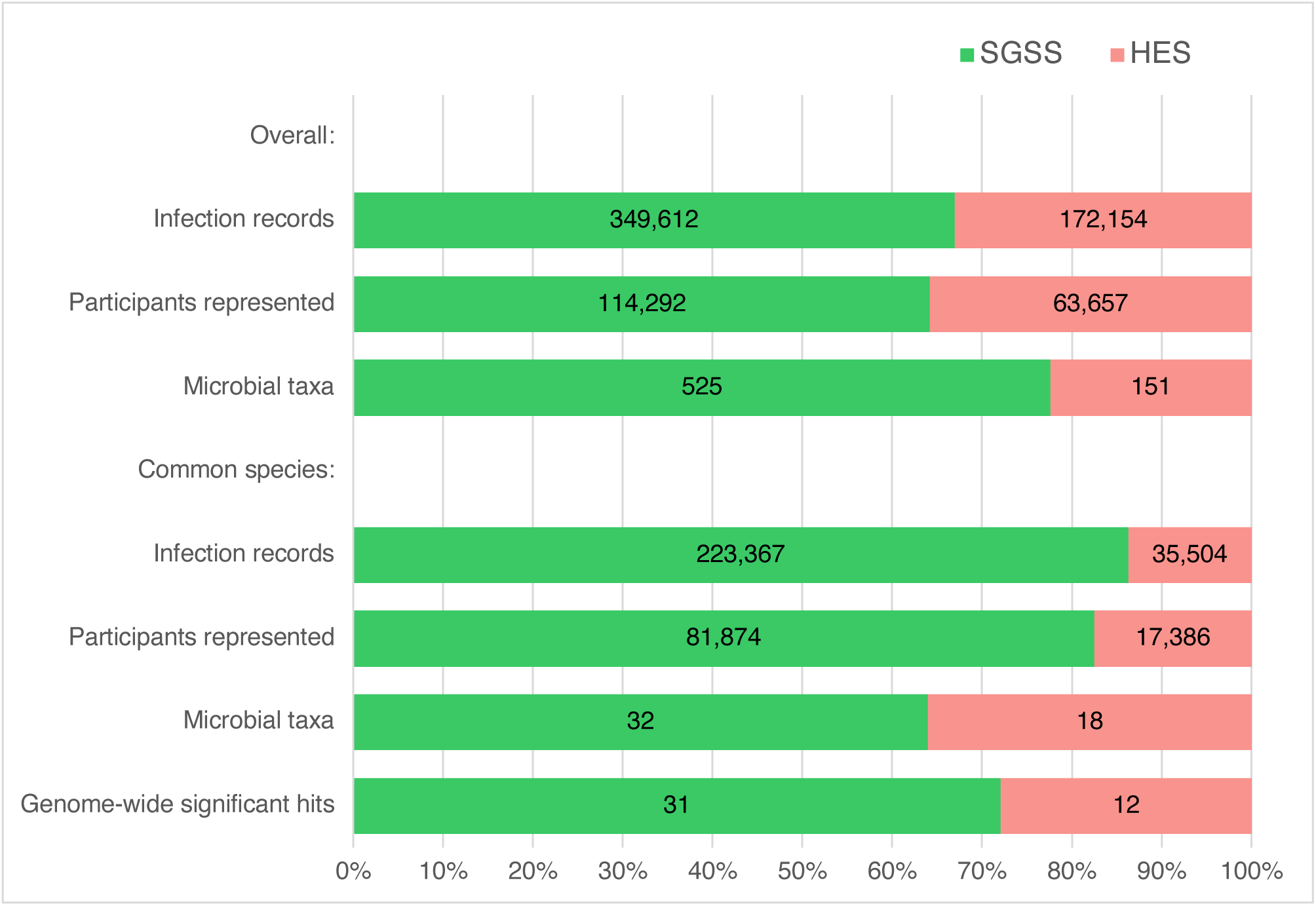
Comparison of infection diagnoses derived from SGSS versus HES linkage to UK Biobank, overall, and restricted to common species in which at least 100 participants are represented. Includes only participants resident in England with complete follow-up.

The infectious organism was available in SGSS from the `ORGANISM SPECIES` field. Usually this identified the microbial taxon at the level of genus or species. For HES, we identified the infectious organism via the disease diagnosis code, defining the narrowest microbial taxon consistent with the three- or four-digit ICD-10 code (Supplementary Table S1). For example, we classified A15.0 “Tuberculosis of lung, confirmed by sputum microscopy with or without culture” as the species *Mycobacterium tuberculosis*, whereas we classified A31.0 “Pulmonary mycobacterial infection”, which most commonly refers to *M. avium*, *M. intracellulare* or *M. kansasii*^42^, simply as the genus *Mycobacterium.* We thereby identified 525 versus 151 distinct microbiological taxa among infection records linked from SGSS versus HES.

### Comparison of common infections in SGSS versus HES

Since the aim was to compare the quality of infection diagnoses for researching underlying risk factors, we focused on infections that were (i) identified at species level (ii) common (i.e., affecting over 100 UKB participants), and (iii) whose genotyping data passed quality control filters (detailed in Methods). Having applied the above criteria, there were 223,367 versus 35,504 infection records from SGSS versus HES (a ratio of 6.3-fold), corresponding to 81,874 versus 17,386 participants (4.7-fold), across 32 versus 17 microbial species (1.8-fold; Figure 1). These comprised 30 versus 8 bacteria, of which 5 were identified in both: *Escherichia coli*, *Haemophilus influenzae*, *Klebsiella pneumoniae*, *Staphylococcus aureus* and *Streptococcus pneumoniae*. Three bacteria were commonly identified through HES only: *Clostridioides difficile*, *Helicobacter pylori* and *Mycobacterium tuberculosis*. Two fungi were commonly identified through SGSS only: *Candida albicans* and *Nakaseomyces glabratus* (formerly *Candida glabrata*). Other pathogens were commonly identified in HES only: eight viruses, including Hepatitis C Virus, Human Papillomavirus and Influenza Virus; and two parasites: *Enterobius vermicularis* and *Plasmodium falciparum*.

Cases exceeded 1,000 participants for 14 species, including seven identified in SGSS only (Table 1): *Citrobacter koseri, Enterococcus faecalis, Enterobacter cloacae, Klebsiella oxytoca* and *Proteus mirabilis;* two identified in HES only (Table 2): *Helicobacter pylori and* Human papillomavirus; and all five identified at species-level in at least 100 participants in both SGSS and HES, mentioned above (Table 3).

**Table 1.**
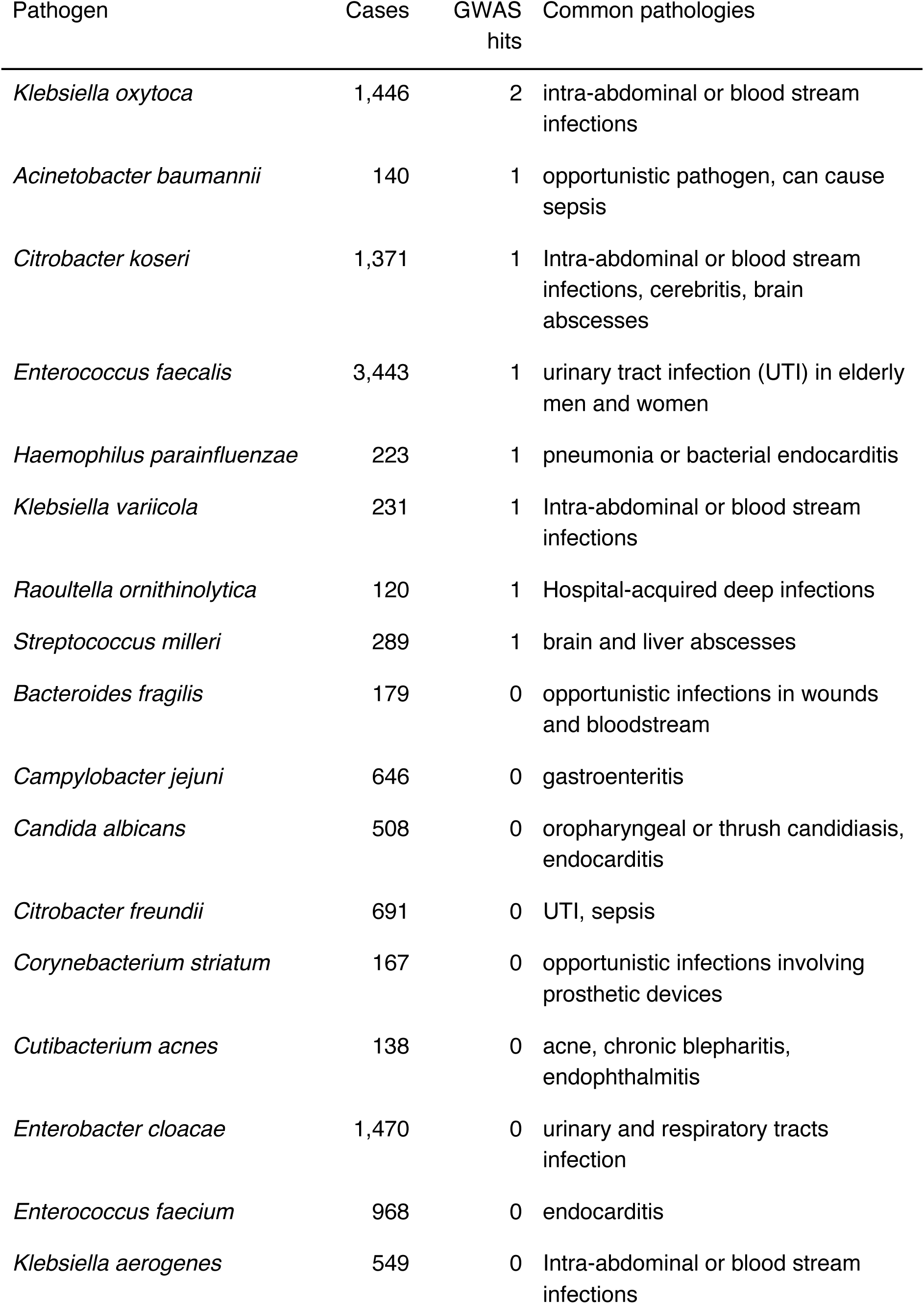
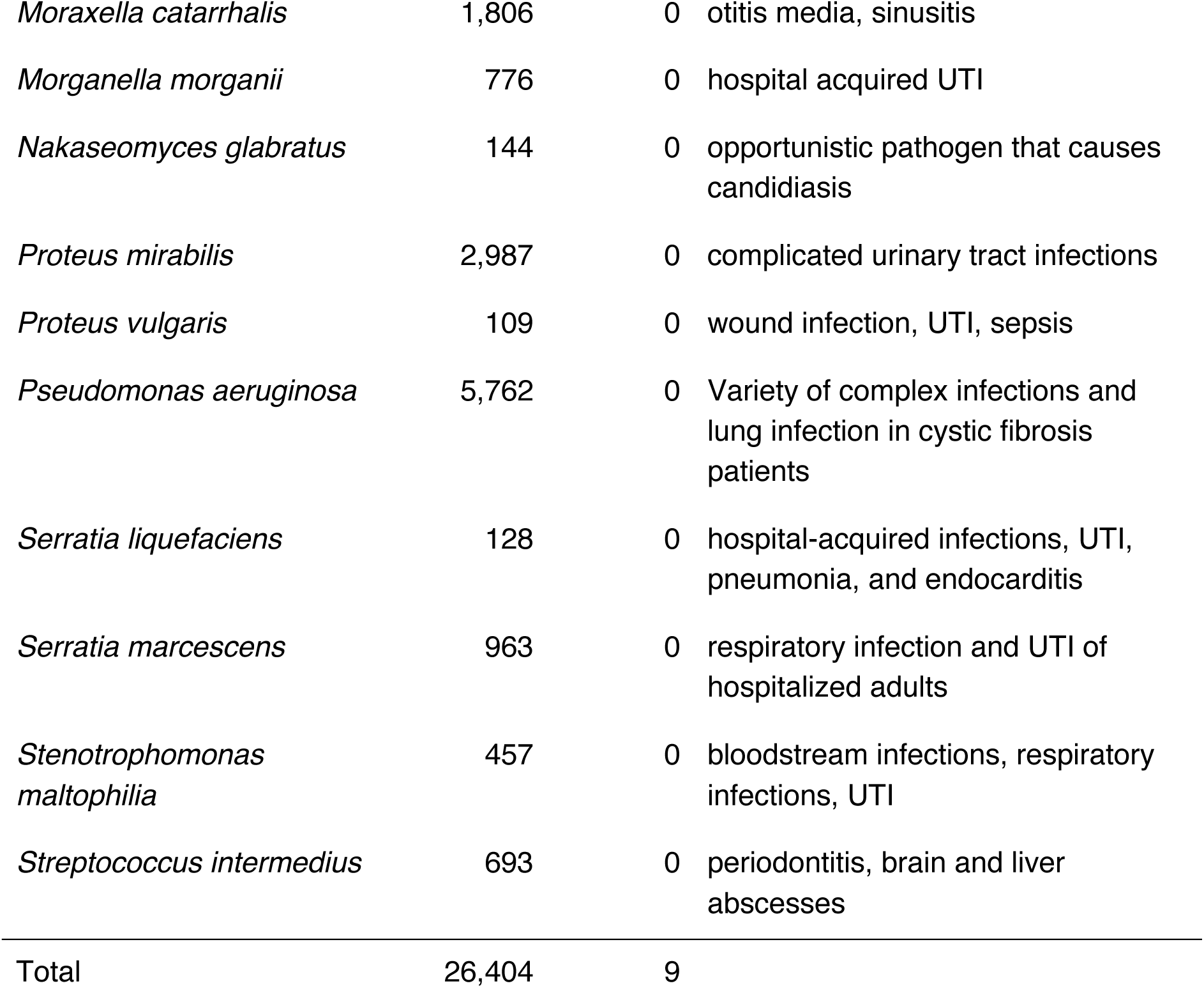
Numbers of cases and genome-wide significant hits for infections commonly identified from UKB linkage to SGSS only.

| Pathogen | Cases | GWAS hits | Common pathologies |
| --- | --- | --- | --- |
| *Klebsiella oxytoca* | 1,446 | 2 | intra-abdominal or blood stream infections |
| *Acinetobacter baumannii* | 140 | 1 | opportunistic pathogen, can cause sepsis |
| *Citrobacter koseri* | 1,371 | 1 | Intra-abdominal or blood stream infections, cerebritis, brain abscesses |
| *Enterococcus faecalis* | 3,443 | 1 | urinary tract infection (UTI) in elderly men and women |
| *Haemophilus parainfluenzae* | 223 | 1 | pneumonia or bacterial endocarditis |
| *Klebsiella variicola* | 231 | 1 | Intra-abdominal or blood stream infections |
| *Raoultella ornithinolytica* | 120 | 1 | Hospital-acquired deep infections |
| *Streptococcus milleri* | 289 | 1 | brain and liver abscesses |
| *Bacteroides fragilis* | 179 | 0 | opportunistic infections in wounds and bloodstream |
| *Campylobacter jejuni* | 646 | 0 | gastroenteritis |
| *Candida albicans* | 508 | 0 | oropharyngeal or thrush candidiasis, endocarditis |
| *Citrobacter freundii* | 691 | 0 | UTI, sepsis |
| *Corynebacterium striatum* | 167 | 0 | opportunistic infections involving prosthetic devices |
| *Cutibacterium acnes* | 138 | 0 | acne, chronic blepharitis, endophthalmitis |
| *Enterobacter cloacae* | 1,470 | 0 | urinary and respiratory tracts infection |
| *Enterococcus faecium* | 968 | 0 | endocarditis |
| *Klebsiella aerogenes* | 549 | 0 | Intra-abdominal or blood stream infections |
| *Moraxella catarrhalis* | 1,806 | 0 | otitis media, sinusitis |
| *Morganella morganii* | 776 | 0 | hospital acquired UTI |
| *Nakaseomyces glabratus* | 144 | 0 | opportunistic pathogen that causes candidiasis |
| *Proteus mirabilis* | 2,987 | 0 | complicated urinary tract infections |
| *Proteus vulgaris* | 109 | 0 | wound infection, UTI, sepsis |
| *Pseudomonas aeruginosa* | 5,762 | 0 | Variety of complex infections and lung infection in cystic fibrosis patients |
| *Serratia liquefaciens* | 128 | 0 | hospital-acquired infections, UTI, pneumonia, and endocarditis |
| *Serratia marcescens* | 963 | 0 | respiratory infection and UTI of hospitalized adults |
| *Stenotrophomonas maltophilia* | 457 | 0 | bloodstream infections, respiratory infections, UTI |
| *Streptococcus intermedius* | 693 | 0 | periodontitis, brain and liver abscesses |
| Total | 26,404 | 9 |  |

**Table 2.** Numbers of cases and genome-wide significant hits for infections commonly identified from UKB linkage to HES only.

| Pathogen | Cases | GWAS hits | Common pathologies |
| --- | --- | --- | --- |
| *Clostridioides difficile* | 869 | 2 | diarrhea, pseudomembranous colitis |
| Human papillomavirus | 1,594 | 2 | genital or anal wart |
| *Enterobius vermicularis* | 140 | 1 | intestine infection, enterobiasis |
| *Helicobacter pylori* | 2,513 | 1 | gastritis, stomach ulcers |
| Human immunodeficiency virus | 176 | 1 | acquired immunodeficiency syndrome |
| *Mycobacterium tuberculosis* | 288 | 1 | respiratory infection, tuberculosis |
| Enterovirus C | 174 | 0 | poliomyelitis |
| Hepatitis C virus | 377 | 0 | chronic liver inflammation,<br>liver cirrhosis |
| Hepatitis B virus | 299 | 0 | chronic liver inflammation,<br>liver cirrhosis |
| Human<br>alphaherpesvirus 3 | 967 | 0 | chickenpox |
| Human<br>orthopneumovirus | 119 | 0 | respiratory tract infection |
| *Plasmodium falciparum* | 149 | 0 | malaria |
| Total | 9,035 | 8 |  |

**Table 3.** Numbers of cases and genome-wide significant hits for infections commonly identified from UKB linkage to SGSS or HES.

| Pathogen | Cases<br>(SGSS) | Cases<br>(HES) | GWAS hits<br>(SGSS) | GWAS hits<br>(HES) | Common pathologies |
| --- | --- | --- | --- | --- | --- |
| *Escherichia coli* | 51,547 | 5,144 | 9 | 1 | opportunistic UTI |
| *Haemophilus*<br>*influenzae* | 5,641 | 489 | 8 | 0 | respiratory tract infections, eye<br>infections, bloodstream<br>infection |
| *Staphylococcus*<br>*aureus* | 20,987 | 3,238 | 4 | 0 | abscesses, respiratory<br>infections, food poisoning, and<br>sepsis |
| *Klebsiella*<br>*pneumoniae* | 4,159 | 844 | 1 | 0 | lung infection, pneumonia |
| *Streptococcus*<br>*pneumoniae* | 1,928 | 631 | 0 | 2 | respiratory tract infections,<br>pneumonia |
| Total | 84,262 | 10,346 | 22 | 3 |  |

To compare the quality of infection diagnoses ascertained through SGSS vs HES for researching risk factors, we counted the number of genome-wide significant associations across the human genome for susceptibility to infection with common pathogens, with a focus on three species: *Escherichia coli*, *Staphylococcus aureus* and *Streptococcus pneumoniae*.

### Case study: *Escherichia coli*

*E. coli* is a common pathogen and gut commensal that causes gastro-intestinal and urinary tract infections. *E. coli* is the commonest cause of blood infection in the UK, with a 15% case fatality rate^43^ and an annual incidence of 150 per 100,000 among those aged 65-74, rising to 430 per 100,000 among those aged 75 and over^44^. Among English-resident UKB participants, 51,547 were identified with an *E. coli* infection from SGSS data compared to 5,144 cases identified from HES data. In SGSS, two-thirds of cases were identified via urine specimens.

In our GWAS, we defined as controls all 234,460 UKB participants who attended English assessment centres, with genotyping data that passed quality control filters, and no infection records, based on SGSS and HES. We followed standard procedures for GWAS, analysing imputed genotypes^45^ with REGENIE^46^, including age and sex as covariates^40^. We excluded genetic variants with low imputation information scores (below 0.3) or low minor allele frequencies (below 0.001). The threshold for genome-wide significance was *p* < 5×10^-8^. To avoid over-counting signals of association, we clustered nearby significant loci (within one megabase) that exhibited high linkage disequilibrium (r^2^ > 0.2), treating them as one signal.

Manhattan plots (Figure 2A) show that one genome-wide significant susceptibility locus predisposing to *E. coli* infection was detected using HES-derived diagnoses, versus nine for SGSS-derived diagnoses (all specimens). Using forest plots (Figure 2B), we compared effect sizes of these nine latter loci between cases defined from HES, SGSS (all specimens) and SGSS (urine specimens). Specifically, we investigated the index variant, defined as the most significant variant at that locus.

**Figure 2.**
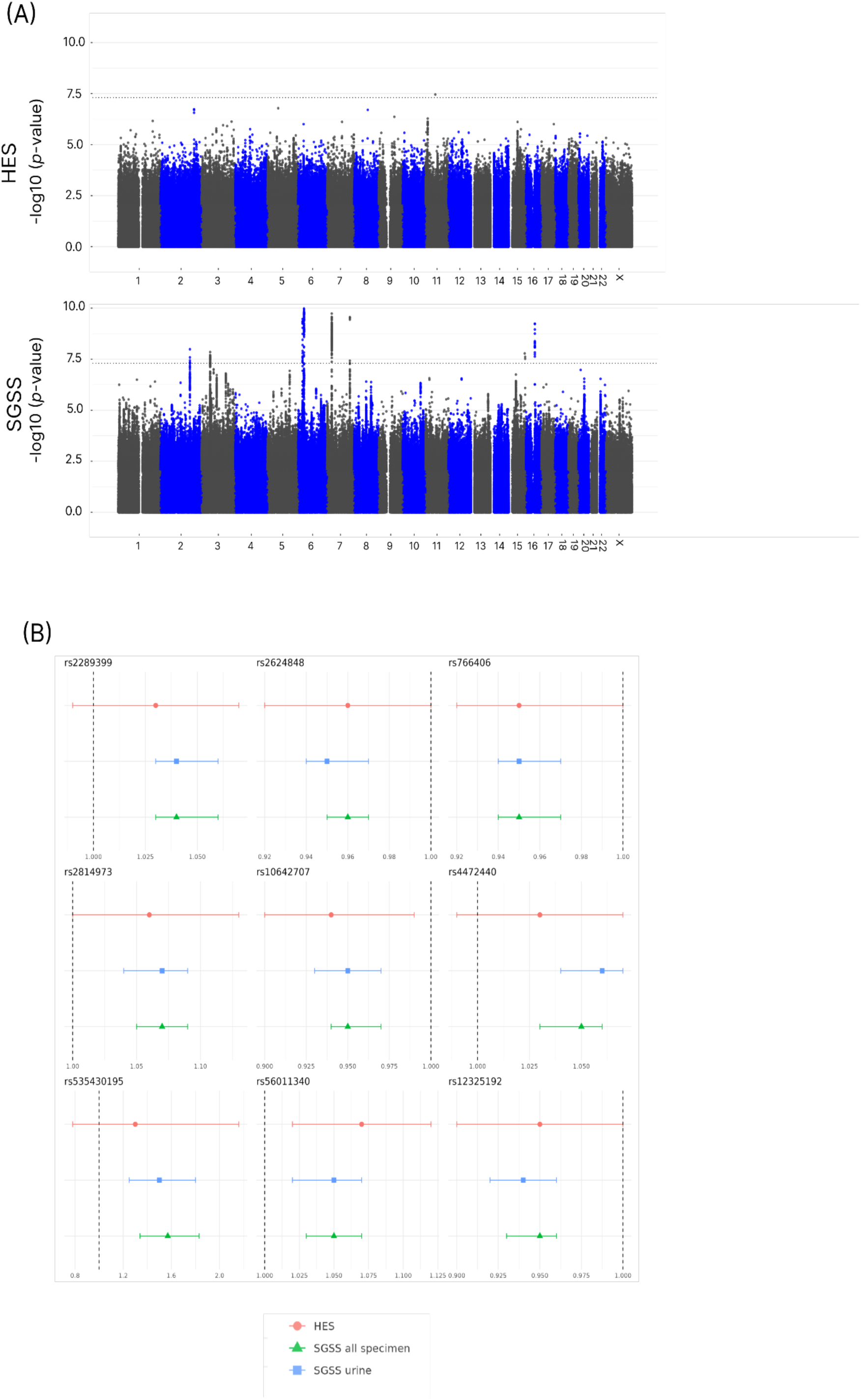
Manhattan plots (A) and forest plots (B) revealing human genetic susceptibility loci to Escherichia coli infection. In (A), black dotted lines represent the genome-wide significance threshold of p = 5 x 10^-8^. In (B), odds ratios of significant E. coli-associated genetic variants identified from SGSS linkage (all specimens) are compared to SGSS urine specimens and HES-linked cases. Error bars represent 95% confidence intervals.

First, we compared the direction of effect between diagnoses derived from SGSS (all specimens) versus HES. The direction of effect represents the sign of the estimated log odds ratio, so a positive effect means that more copies of the alternative allele at an index variant were associated with increased susceptibility to infection, and vice versa. For all nine index variants, the direction of effect was consistent between HES-derived diagnoses and SGSS-derived diagnoses (all specimens). This is compatible with an interpretation that the discrepancy in statistical significance is explained by smaller case numbers, causing wider confidence intervals, in diagnoses derived from HES versus SGSS (all specimens). Indeed, only six of the index variants (rs2624848, rs766406, rs2814973, rs10642707, rs56011340, and rs12325192) were nominally significant at the 5% level for the HES-derived diagnoses.

All nine variants were nominally significant at the 5% level using diagnoses derived from SGSS urine specimens, and six were genome-wide significant. The exceptions were rs2289399, rs535430195 and rs56011340, yielding *p*-values of 7.2 x 10^-7^, 2.0 x 10^-5^ and 1.9 x 10^-5^ respectively. Again, this is consistent with a reduction in statistical power owing to smaller sample size.

We did not further investigate the index variant of the genome-wide significant locus identified from HES data because of concerns over genotyping quality. The index variant rs573670460 (*p*-value 3.5 x 10^-8^) only marginally passed our quality control criteria in terms of minor allele frequency (0.002) and imputation information score (0.51), while the Manhattan plot revealed an isolated variant lacking the typical peak of nearby genome-wide significant variants expected due to local linkage disequilibrium. These factors suggest it may have been a spurious association.

Of course, a major limitation of our study’s approach – counting the number of significant associations to assess the quality of diagnoses for researching infection-related risk factors – is that we do not know whether each signal is genuine or artefactual. To mitigate this concern, we considered the biological plausibility of the index variant at each significant susceptibility locus. Eight of the nine genome-wide significant index variants reside within protein-coding genes. Using the NHGRI-EBI human GWAS catalog, several appear to be involved in biologically relevant pathways like immune response or urogenital function^47–59^ (Supplementary Table S2). Together with the robustness of genetic associations to mediation, careful control of confounding by standard GWAS methodology, and the stringency of the threshold for genome-wide significance, this supports the interpretation that SGSS data contribute substantial scientific added value over HES data for studying *E. coli* infection in UK Biobank.

### Case study: *Staphylococcus aureus*

*S. aureus* is a major pathogen and the second commonest cause of blood infection in the UK, inflicting a 22% case fatality rate^43^, with an annual incidence of 21 per 100,000 across all age groups, rising by more double among men aged 65-74 and women aged 75 and over, and more than quadruple among men aged 75 and over^60^. *S. aureus* commonly colonizes the nose and skin, and frequently causes skin, soft tissue, and other infections^61^. Among English-resident UKB participants, 20,987 were diagnosed with a *S. aureus* infection through SGSS data compared to 3,228 identified by HES data. The dominant specimen types in SGSS were nose swabs (4,229 cases), skin wounds (3,620 cases), and skin trauma (2,535 cases).

Four genome-wide significant loci predisposing to *S. aureus* infection were detected using SGSS-derived diagnoses (all specimens): rs9271494, rs375309052, rs9267947 and rs529250272 (Supplementary Table S3). No loci were genome-wide significant using HES-derived diagnoses (Figure 3A). Forest plots (Figure 3B) show that the effect sizes of these four loci were consistent in direction, and similar in magnitude, whether cases were defined from HES or SGSS, and regardless of specimen type.

**Figure 3.**
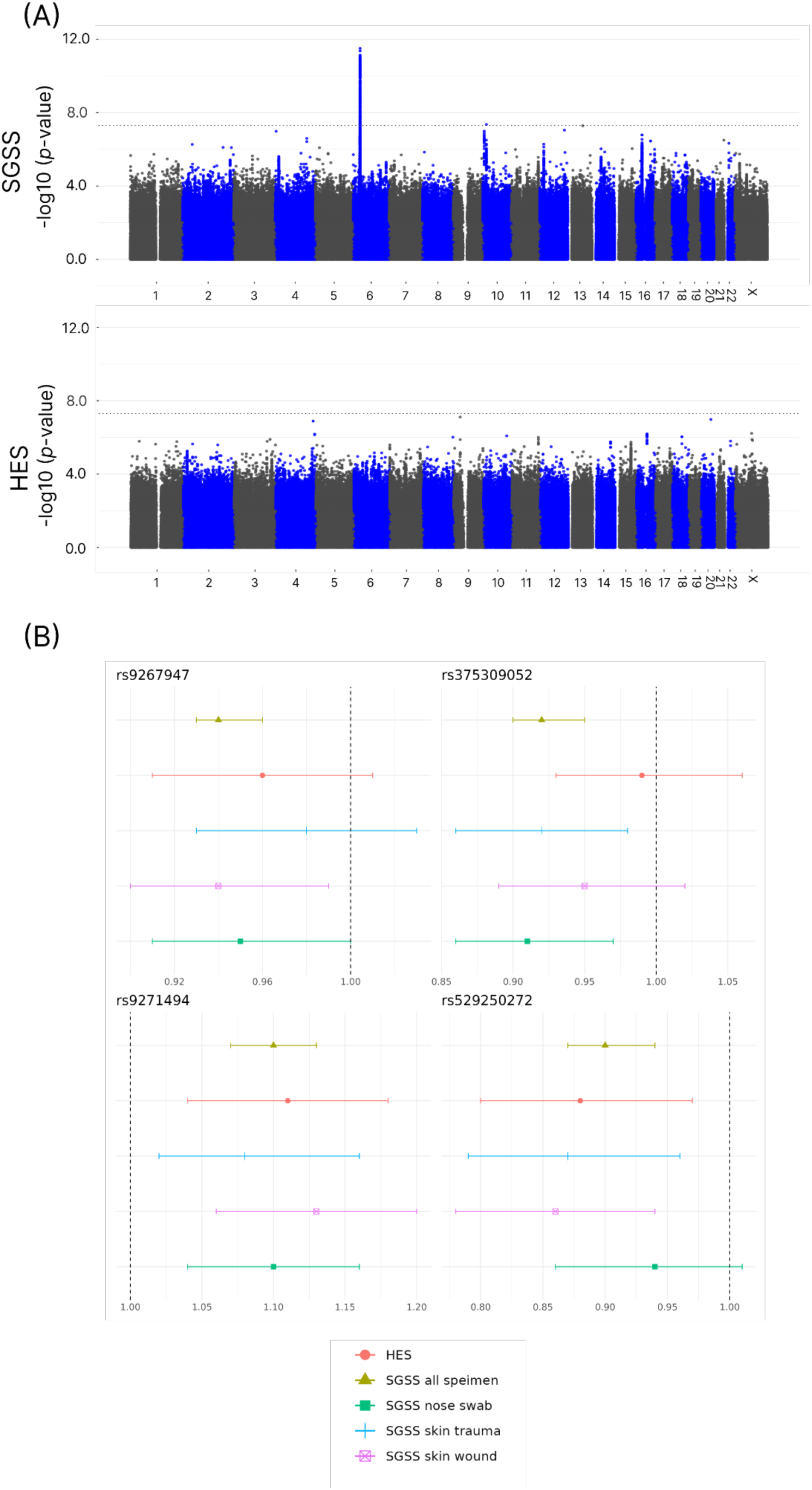
Manhattan plots (A) and forest plots (B) revealing human genetic susceptibility loci to Staphylococcus aureus infection. In (A), black dotted lines represent the genome-wide significance threshold of p = 5 x 10^-8^. In (B), odds ratios of significant S. aureus-associated genetic variants identified from SGSS linkage (all specimens) are compared to SGSS other specimens and HES-linked cases. Error bars represent 95% confidence intervals.

Looking at nominal significance, three of the four index variants were significant at the 5% level when restricting attention to the SGSS nose swab (rs9267947, rs375309052, and rs9271494), skin wound (rs375309052, rs9271494, and rs529250272), and skin trauma (rs375309052, rs9271494, and rs529250272) specimen-based case definitions. Two of the four (rs9271494 and rs529250272) were also nominally significant using HES-derived diagnoses.

In the context of evaluating the added value of SGSS versus HES data for analysing determinants of microbiological infection, we briefly considered the biological plausibility of these associations. Two of the four genome-wide significant index variants reside within protein-coding genes, while three of the four fell within the major histocompatibility complex^62–65^ (Supplementary Table S3). *S. aureus* infection is one of the few bacterial infections with susceptibility loci listed in the GWAS catalog^47^. Twelve genome-wide significant variants (*p* ≤ 1×10⁻⁶) have reported associations with phenotypes associated with susceptibility to *S. aureus* infection including “*Staphylococcus aureus* infection” and “Skin and soft tissue *Staphylococcus aureus* infection” (Supplementary Table S4). Two of those (rs115740542 and rs4321864) showed nominal significance for diagnoses derived from SGSS all specimens, yielding *p*-values of 1.5×10⁻⁴ and 4.1×10⁻⁴, respectively^66–69^, supporting the view that SGSS data produces meaningful signals for the study of *S. aureus* infection in UK Biobank.

### Case study: *Streptococcus pneumoniae*

*S. pneumoniae* is an important pathogen causing a range of diseases, notably pneumonia, meningitis and blood infection, and is commonly carried in the nose and throat^72,73^. Among English-resident UKB participants, 1,928 were diagnosed with *S. pneumoniae* infection from SGSS data compared to 631 identified from HES data. Over 70% of cases were identified via sputum.

In our analyses, *S. pneumoniae* infection was unusual in that no genome-wide significant susceptibility loci were detected using SGSS-derived diagnoses, whereas two loci were detected using HES-derived diagnoses (Figure 4A, Supplementary Table S5, Supplementary Table S6), despite 3.1-fold fewer cases in the latter. The forest plots for the index variants rs61994223 and 13:113432374_TC_T (Figure 4B) differed from the pattern seen in *E. coli* and *S. aureus*. Although the direction of effect was consistent between cases derived from HES and SGSS (all specimens), the 95% confidence intervals did not overlap, with the genetic risk substantially weaker for cases derived from SGSS. The 95% confidence interval for the latter was narrow and failed to exclude the null hypothesis of no genetic risk.

**Figure 4.**
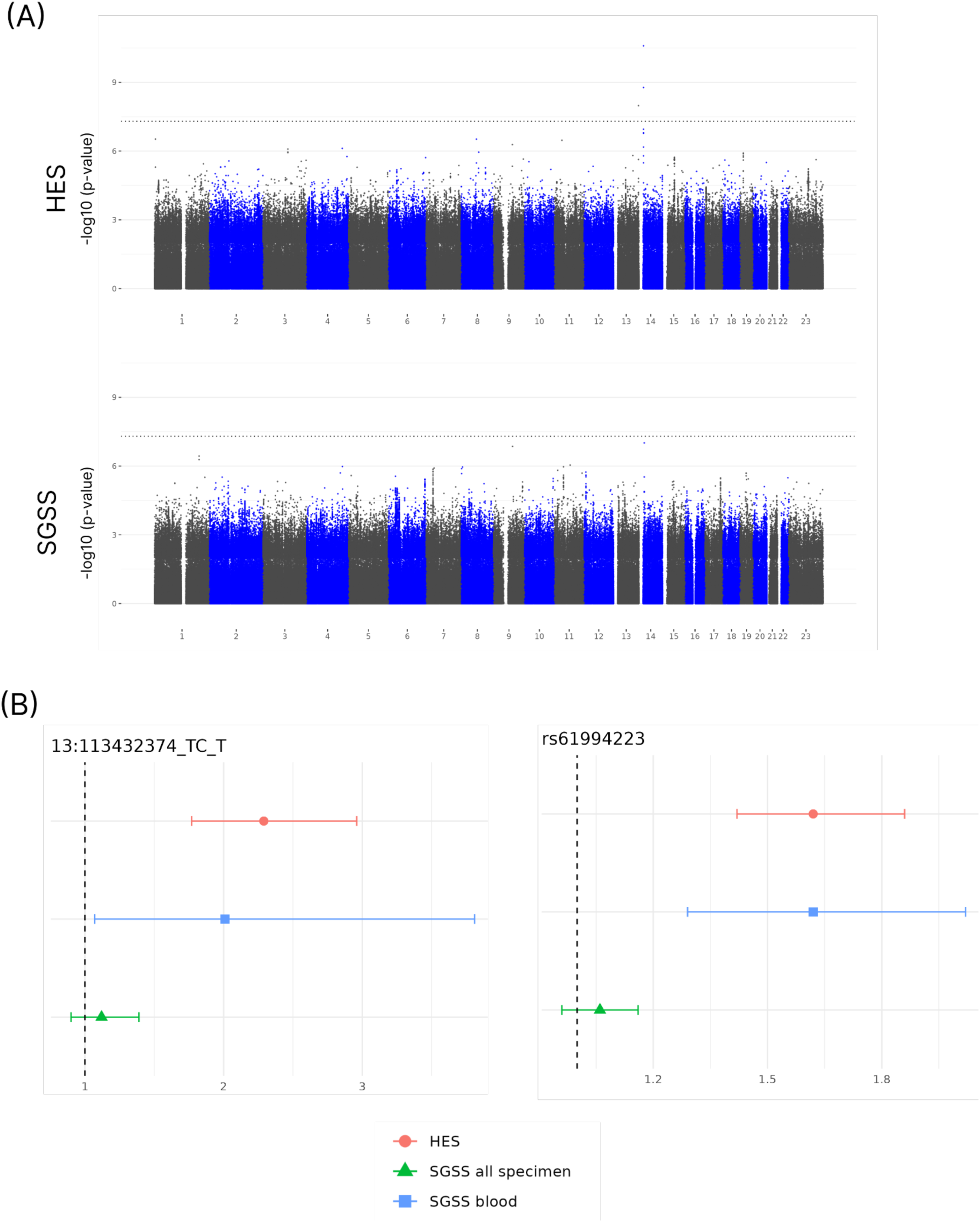
Manhattan plots (A) and forest plots (B) revealing human genetic susceptibility loci to Streptococcus pneumoniae infection. In (A), black dotted lines represent the genome-wide significance threshold of p = 5 x 10^-8^. In (B), odds ratios of significant S. pneumoniae-associated genetic variants identified from SGSS linkage (all specimens) are compared to SGSS other specimens and HES-linked cases. Error bars represent 95% confidence intervals.

Next we used the forest plots to compare the genetic risk in cases derived from SGSS blood specimens (Figure 4B). While the confidence intervals were larger (as expected from smaller case numbers), for both index variants they were consistent with the odds ratios estimated from HES-derived cases; the point estimates were closer to those estimated from HES than SGSS all specimens; and the 95% confidence intervals excluded the null hypothesis of no genetic risk.

These patterns are compatible with an interpretation that HES-derived cases were enriched for more severe *S. pneumoniae* infections, and those infections had a genetic risk profile similar to the blood infections identified from SGSS. In support of this interpretation, we found that only 239 cases of *S. pneumoniae* infection identified from SGSS (all specimens) were also identified from HES, of which 51 had the ICD-10 code A40.3, “Sepsis due to *S. pneumoniae*” (Supplementary Table S5). Past GWAS results suggest that the significant loci identified from HES-derived cases, both of which reside in protein-coding genes, are involved in antimicrobial response^74–80^_(Supplementary Table S6).

### Genome-wide significant associations in SGSS versus HES

To compare the quality of infection diagnoses for researching risk factors more broadly, we counted the number of genome-wide significant associations across the human genome for susceptibility to infection with all 45 common pathogens, diagnosed from SGSS (32 species) or HES (17 species). Five pathogens were analysed twice, using SGSS and HES, making for 49 analyses in total.

In total, and incorporating the case studies detailed above, we found 31 versus 11 genome-wide significant associations across pathogens using SGSS versus HES linkage (Table 1-Table 3).

None of these 42 signals of association overlapped. Half (22/42) of the significant loci predisposed to infection with the three species most commonly diagnosed using SGSS or HES: *Escherichia coli* (56,691 cases, 10 loci), *Haemophilus influenzae* (6,130 cases, 8 loci) and *Staphylococcus aureus* (24,225 cases, 4 loci), perhaps reflecting better statistical power. However, not all well-represented pathogens yielded significant susceptibility loci, for example *Pseudomonas aeruginosa* (5,762 cases).

Of the species commonly identified in both SGSS and HES (Table 3), the significant susceptibility loci for *E. coli*, *H. influenzae* and *S. aureus* infection were mainly (21/22) discovered via SGSS, which accounted for 8.8-fold more diagnoses (78,175 versus 8,871). A further susceptibility locus was discovered via SGSS for *Klebsiella pneumoniae* infection. As mentioned above, *Streptococcus pneumoniae* infection bucked the trend, with both significant susceptibility loci discovered via HES, despite 3.1-fold more diagnoses in SGSS (1,928 versus 631).

Among the species commonly identified in either SGSS or HES, but not both, 17 significant susceptibility loci were identified, nine for each (Table 1-Table 2). Notably, this included four loci predisposing to infection with the three bacteria commonly diagnosed via HES but not SGSS: *Clostridioides difficile* (869 cases, 2 loci), *Helicobacter pylori* (2,513 cases, 1 locus) and *Mycobacterium tuberculosis* (288 cases, 1 locus).

The remaining susceptibility loci predisposed to SGSS-diagnosed infection with *Acinetobacter baumannii* (140 cases), *Citrobacter koseri* (1,371 cases), *Enterococcus faecalis* (3,443 cases), *Haemophilus parainfluenzae* (223 cases), *Klebsiella oxytoca* (1,446 cases, 2 loci), *Klebsiella variicola* (231 cases), *Raoultella ornithinolytica* (120 cases) and *Streptococcus milleri* (289 cases), and HES-diagnosed infection with *Enterobius vermicularis* (140 cases), HIV (176 cases)and Human Papillomavirus (1,594 cases, 2 loci).

### Stratifying by specimen type in SGSS

Infection records linked from SGSS included a SPECIMEN TYPE field, of which the commonest entries for UKB participants were urine/kidney (2.80%), skin/wound (0.57%), sputum (0.41%) and blood (0.16%, taking 2015 as an example^7^). This information affords an opportunity to stratify microbiological diagnoses by site of infection or, more precisely, site of isolation. To some extent, specimen type serves as a proxy for severity of infection, for example allowing us to infer an *E. coli* blood infection rather than a urinary tract infection. In practice, one would like to understand whether a risk factor was related to *E. coli* infection generally, or urinary tract infection specifically, particularly since the large number of urine/kidney isolates could dominate any significant signals of association.

We investigated stratifying SGSS-derived infection diagnoses by specimen type by comparing *all specimen* types, described above, to the *dominant specimen* types, i.e. the most numerous specimen type per species. As before, we counted the number of genome-wide significant associations across the human genome for susceptibility to infection (Table 4).

**Table 4.** Numbers of genome-wide significant hits for infections commonly identified from UKB linkage to SGSS: comparison of all-specimen vs dominant-specimen analyses.

| Pathogen | Signals<br>in both | All-specimen<br>only | Dominant-specimen<br>only | Dominant<br>specimen<br>type | # of cases (all-specimen / dominant-specimen) |
| --- | --- | --- | --- | --- | --- |
| *Haemophilus influenzae* | 8 | 0 | 0 | Sputum | 5641 / 5160 |
| *Escherichia coli* | 6 | 3 | 2 | Urine | 51547 / 34860 |
| *Staphylococcus aureus* | 0 | 4 | 1 | Nose swab | 20987 / 4229 |
| *Klebsiella oxytoca* | 0 | 2 | 1 | Urine | 1446 / 565 |
| *Citrobacter koseri* | 0 | 1 | 0 | Urine | 1371 / 637 |
| *Enterococcus faecalis* | 0 | 1 | 0 | Urine | 3443 / 1766 |
| *Haemophilus parainfluenzae* | 0 | 1 | 0 | Sputum | 223 / 161 |
| *Klebsiella pneumoniae* | 0 | 1 | 0 | Urine | 4159 / 2064 |
| *Proteus mirabilis* | 0 | 0 | 1 | Urine | 2987 / 1668 |
| Total | 14 | 13 | 5 |  |  |

In total, incorporating the case studies detailed previously, we found 32 significant associations with susceptibility to infection across 10 species based on SGSS-derived diagnoses considering all specimen types, dominant specimen types, or both. Five of these were detected only when considering dominant specimen types, 13 were detected only when considering all specimen types, and 14 were detected either way. The five susceptibility loci detected only when considering dominant specimen types comprised two for *Escherichia coli* infection (urine), and one each for *Staphylococcus aureus* infection (nose swab), *Proteus mirabilis* infection (urine) and *Klebsiella oxytoca* infection (urine).

## Discussion

In this study, we evaluated whether SGSS data could enhance infection research in UK Biobank by providing more numerous and more specific case definitions of infection events than HES data.

After quality control, 6.8-fold more infection records (223,367 versus 32,952) were derived from SGSS versus HES linkage among English-resident UKB participants. Deduplicating by participant, SGSS yielded 5.0-fold more cases of infection than HES (81,874 versus 16,345). SGSS records provided additional details like site of microbiological isolation and the cultured organism, usually at species-level. Focusing on common species infecting over 100 English-resident UKB participants, SGSS facilitated analysis of 1.9-fold more pathogens than HES (32 versus 17).

There were notable exceptions to these trends. Most viral pathogens were identified predominantly via HES, including all eight common species that numbered over 100 cases. SGSS records were not uniformly more abundant than HES, even for studying bacterial infections. The important pathogens *C. difficile*, *H. pylori* and *M. tuberculosis* were identified more commonly from HES than from the SGSS AMR table that we focused on here, which primarily contains laboratory-confirmed bacterial infections based on culture. Fewer than 100 cases of each were identified in the AMR table. Like viruses, these bacteria are commonly diagnosed using methods other than microbiological culture, including PCR, enzyme immunoassays, breath tests, endoscopy, and radiological examination, often in conjunction with clinical symptoms. For tuberculosis, more cases may appear in the SGSS CDR (Communicable Disease Reporting) table, which holds statutory notifications of notifiable diseases in England, but these notifications are not necessarily based on microbiological confirmation and were not the focus of our analysis.

To assess the quality of SGSS- and HES-derived case definitions for researching infection-related risk factors, we counted the number of loci in the human genome significantly associated with susceptibility to common infections. Analysing genetic risk factors helped us avoid issues surrounding mediation and mitigated against confounding using standard GWAS approaches.

SGSS revealed 2.6-fold more significant susceptibility loci than HES (31 versus 12). Of the species commonly identified in both SGSS and HES, the significant susceptibility loci for *E. coli*, *H. influenzae* and *S. aureus* infection were mainly (21/22) discovered via SGSS, which accounted for 8.8-fold more diagnoses (78,175 versus 8,871). The overarching trend was for larger sample sizes to yield greater numbers of significant susceptibility loci, based on comparisons of case numbers between HES and SGSS, and between specimen types within SGSS.

Yet even when SGSS yielded more cases than HES, it did not always yield more significant susceptibility loci. In the case of *S. pneumoniae* infection, SGSS yielded 3.1-fold more cases than HES (1,928 versus 631), but analysis of the HES-derived cases revealed two significant susceptibility loci. Stratifying the SGSS-derived cases by specimen type suggested the presence of heterogeneity in risk profile between blood infections, representing the most severe disease, and sputum infections, representing the most numerous specimen type. SGSS-derived blood infections showed a similar genetic risk profile to HES-derived infections. Heterogeneity in genetic susceptibility to different clinical manifestations of infection was also evident for *E. coli*, where the index variant at the *CALD1* locus showed a substantially larger odds ratio in the context of SGSS-derived urine specimens than all specimens. Interestingly, *CALD1* is involved in smooth muscle contraction, which has been implicated particularly in urinary tract infections^55,56^. These examples suggest that the benefit, in terms of larger sample size, of pooling specimen types in SGSS is sometimes offset by diluting the specificity of case definitions, with neither approach beating the other in all situations.

These patterns, both the broad trends and their exceptions, reflect the distinct characteristics of SGSS and HES infection records. HES was conceived as a system to collect data on hospital activity for administration and clinical audit, whereas SGSS was created to centralize data from microbiology laboratories and practitioners for monitoring infection, outbreaks and antimicrobial resistance. Differences in diagnosis reflect hospital clinician-centred versus microbiology laboratory-centred processes, with the former coded post-discharge using ICD-10 based on patient notes, and the latter coded taxonomically, usually at species level. The events that lead to HES versus SGSS infection records differ: HES records are mainly precipitated by hospital stays in which an infection diagnosis was made, whereas SGSS records reflect microbiological culture of specimens sent from hospitals or general practices suspecting infection. These differences and others generate contrasting profiles of patients, infections, disease severities, and recording practices.

There were numerous limitations to our study. (1) We concentrated on the SGSS AMR table, which forms the bulk of SGSS data, but we omitted the CDR, which records notifiable diseases and other surveillance priorities. Our SGSS analyses therefore ignored certain pathogens in SGSS not subjected to culture-based tests, particularly viral infections. (2) In quantifying the utility of SGSS or HES-derived diagnoses for infection research, we counted the number of significant susceptibility loci. Usually we do not know whether a given genetic variant genuinely predisposes to infection, or not. To mitigate this, we considered the biological plausibility of significant associations for exemplar organisms, but our approach was limited in scope and based on existing literature. (3) We performed GWAS on pathogens identified at the species level, assuming that species-level resolution provides a superior basis for understanding the aetiology of infection. However, pooling diseases can also reveal insights^81^, an aspect of the SGSS versus HES comparison that we overlooked by discarding coarser taxonomic diagnoses. (4) To compare SGSS and HES, it was necessary to convert ICD-10 codes into taxonomic groups, but this process inevitably introduces idiosyncrasies and inaccuracies. (5) For most species, the sample sizes were modest, particularly when stratifying infections by specimen type to identify the most severe pathologies. Although case numbers can only increase in the future, the need to focus on currently well-represented species limits the generalizability of our conclusions.

In summary, we have demonstrated that linking SGSS records to UKB adds significant value to infection studies by substantially increasing both the number of infection cases and the variety of pathogens captured. We have characterized the strengths of SGSS and HES record linkage for understanding infection, highlighting contrasting profiles of patients, infections, disease severities and recording practices. This presents opportunities to gain further insights by integrating information across these and other databases, because, of course, in practice, they would not be used mutually exclusively. To conclude, SGSS offers a valuable source of data for investigating risk factors for infection in UKB, with the potential to reveal new insights into both genetic and non-genetic risk factors, as well as the role of infection as an exposure for other, non-communicable, diseases.

## Methods

### Record linkage

As previously described^7,36^, we used OpenPseudonymiser to compare pseudonymised (tokenised) NHS numbers present on SGSS records with tokenised NHS numbers of UKB participants. We exported the following fields from the SGSS AMR table: SPECIMEN_DATE, SPECIMEN_TYPE_DESC, `SPECIMEN_GROUP_DESC`, `ORGANISM_GENUS_NAME`, `ORGANISM_SPECIES_NAME`, `ORGANISM_SUBSPECIES_NAME`, `LAB_NAME`, using the UKB EID as the primary key. We considered isolates received from English-resident UKB participants until 12^th^ November 2021. For comparison, the latest records analysed from HES were dated 7^th^ May 2021. The earliest records analysed for SGSS and HES respectively were 10^th^ October 2000 and 2^nd^ December 1980; however, very few records were dated so early. More representative start dates for SGSS specimens and HES episodes respectively, covering 99.9% of records, were 3^rd^ December 2008 and 8^th^ April 1997.

### Inclusion criteria for UK Biobank participants

The aim of the GWAS was to evaluate the added value of linking SGSS-derived infection records to UKB for research purposes. Since SGSS records for Wales and Scotland are incomplete, we focused on England. To begin, we considered all UKB participants with available whole-genome genotyping, based on Genome Reference Consortium Human Build 37 (GRCh37), for inclusion in our GWAS analysis. We applied several exclusion criteria to refine the study cohort (Supplementary Table S7). First, we excluded participants who were lost to follow-up, either due to withdrawal of consent or because the NHS lost contact without an official death certificate being recorded by the Death Registry. Second, we excluded non-English residents as determined by the UKB assessment centre. Finally, we implemented genotyping quality control measures: we excluded participants with atypical sex chromosome configurations (neither XX nor XY), those whose reported sex did not match the sex inferred from genotype data, and individuals who were outliers in heterozygosity and missing genotype rates^45^. We also removed samples not included in UKB’s kinship inference procedure or genotype phasing process, as well as those with more than ten putative third-degree relatives among the participants, based on estimates of excess relatedness^45^.

### Case and control definitions

We defined infection cases in UKB participants using SGSS- and HES-derived records. In SGSS, a case was identified by any record linking the culture of a specific organism identified at species-level resolution to a participant. In HES, a case was identified by any record linking an ICD-10 diagnosis code mappable to a pathogen at species-level resolution (Supplementary Table S1) to a participant. Where a participant was linked to multiple infection records concerning the same pathogen (and same specimen type in the case of SGSS linkage), we considered only the first occurrence. In defining controls, we excluded any participant with any infection record in either HES, SGSS, or both, resulting in a common pool of 234,460 controls for every GWAS.

### GWAS

To maintain adequate statistical power, our GWAS focused on pathogens with over 100 cases. Using the imputed genetic data from UKB^45^, we performed GWAS using REGENIE^46^, with age and sex as covariates. When interpreting the GWAS results, we filtered out any genetic variant with an imputation information score below 0.3 or minor allele frequency below 0.001. We employed the conventional genome-wide significance threshold of *p* < 5×10^-8^.

We utilized a clumping method to identify independent genetic association signals while accounting for linkage disequilibrium between variants. To begin, this process^82^ ranked all variants by *p*-value from most to least significant. Starting with the most significant variant, designated as the index variant, we identified all other variants within one megabase in linkage disequilibrium with the index variant at a level exceeding r^2^ = 0.2. These variants were “clumped” with the index variant, and considered as a single locus. The clumped variants were excluded from downstream analyses to avoid redundancy. We repeated this procedure iteratively, selecting the next most significant variant not already clumped, and applied the same criteria until all significant variants had been evaluated.

### Ethics statement

UKHSA gathers data from NHS microbiology laboratories, storing it in the SGSS database. That data is fully identified, and anonymised extracts are generated prior to routine epidemiological analysis, an activity permitted under Section 251 of the National Health Service Act 2006, which allows processing of named patient data without consent for defined purposes, including public health surveillance. Participants in UK Biobank gave written, informed consent for UK Biobank to follow their health using linkage to electronic health-related records. All methods were carried out in accordance with relevant guidelines and regulations as detailed in the ethical framework used by UK Biobank^83^. This work was reviewed by both the UK Biobank Ethical Committee and the UKHSA Caldicott Guardian. No experimental protocols requiring prior institutional and/or licencing committee approval were undertaken.

## Data Availability

The code written for database linkage in this study is internal to UK Health Security Agency (UKHSA) systems, and will not be released publicly. It is planned that data provided by UKHSA will be incorporated into the UK Biobank database and released through the usual UK Biobank governance processes. Work to effect this is ongoing, and UK Biobank registered researchers will be notified in the normal way.

https://www.ukbiobank.ac.uk/enable-your-research/register

## Acknowledgements

This research has been conducted using the UK Biobank Resource under Application Number 53100. We acknowledge the support of the UK Biobank participants, the UK Biobank staff and members of the National Infection Service Information Management team. We thank Chris Spencer and David Eyre for helpful comments.

## Author contributions

Conceptualization: NA, AJM, DHW, DJW. Funding acquisition: NA, DHW, DJW. Data curation, software, analysis: SKL, JA, GBL. Supervision: DHW, DJW. Project administration: NA, AJM, DHW, DJW. Drafting: SKL, DJW. Writing: all authors.

*Authors’ note*: DHW jointly led this work, he secured seed funding, supervised, reviewed and edited earlier drafts of the work, and his writing contributed to the final paper, but he passed away before the work was finalized.

---

Supplementary Table S1: Microbial taxa inferred from three- or four-digit ICD-10 codes. Note that in text we have used the convention of preceding the fourth digit with a decimal point, whereas in the table we have omitted the decimal places.

**Supplementary Table S2:**
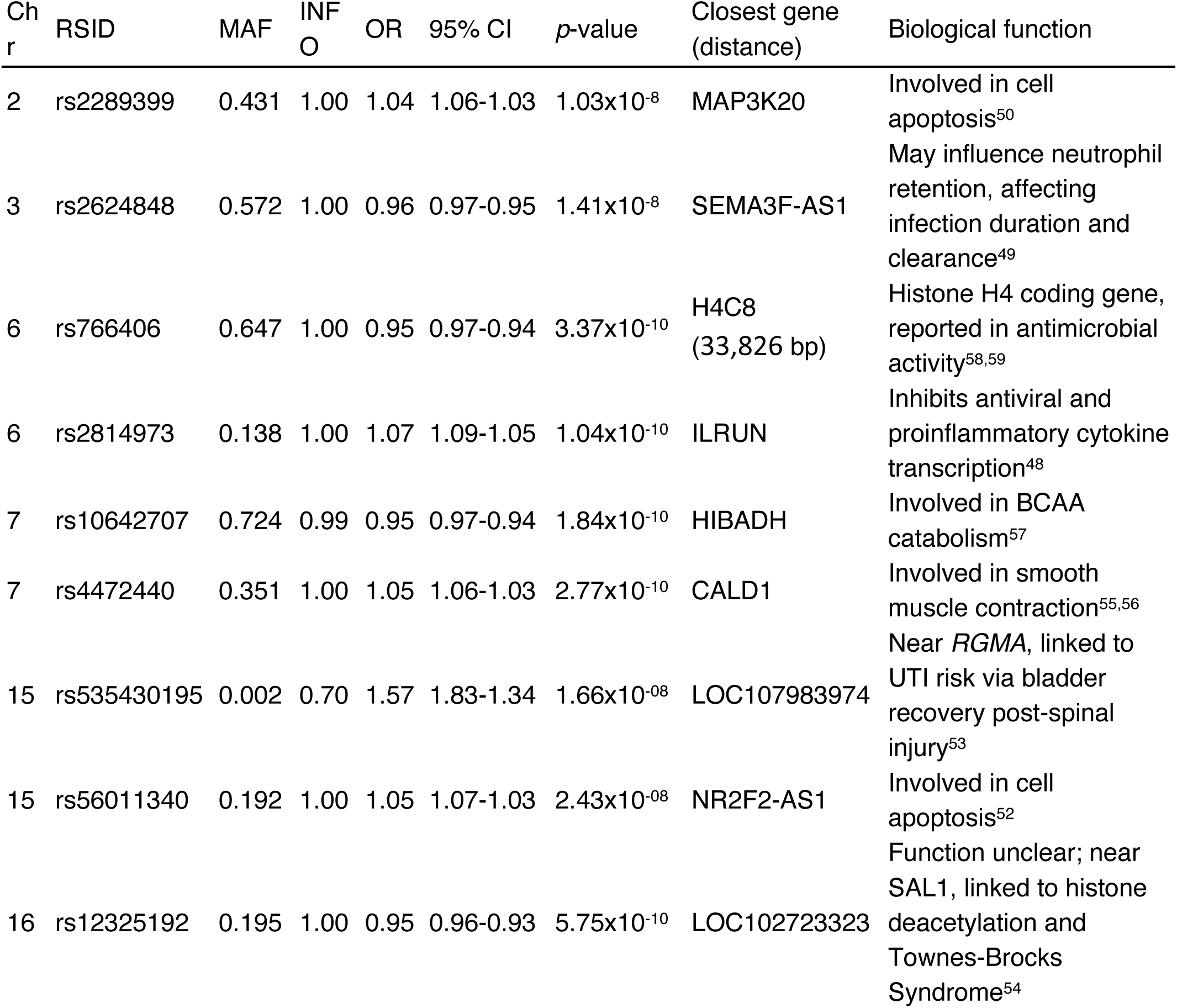
Genome-wide significant susceptibility loci for *E. coli* infection, identified by SGSS linkage. Chr: chromosome; RSID: reference SNP cluster ID; MAF: minor allele frequency; INFO: imputation information score; OR: odds ratio; CI: confidence interval.

**Supplementary Table S3:**
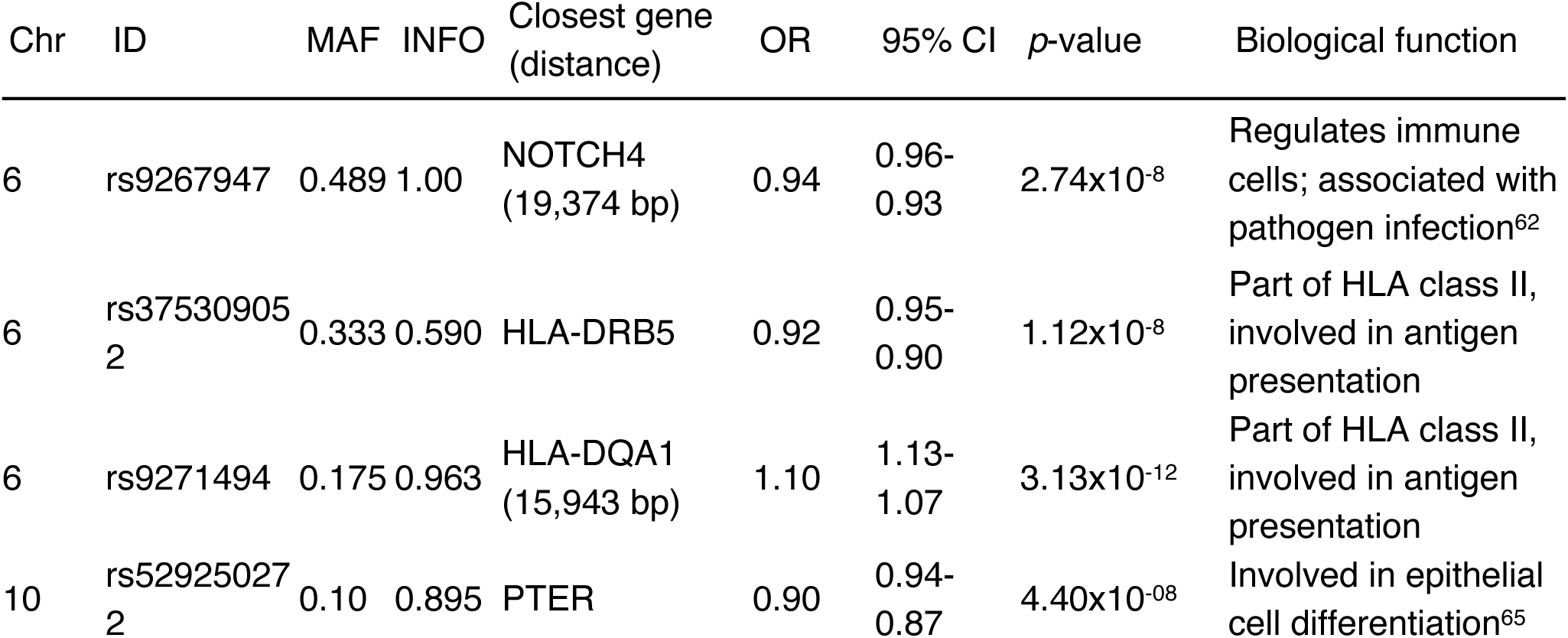
Genome-wide significant susceptibility loci for *S. aureus* infection, identified by SGSS linkage. Chr: chromosome; RSID: reference SNP cluster ID; MAF: minor allele frequency; INFO: imputation information score; OR: odds ratio; CI: confidence interval.

Supplementary Table S4: Comparison of significant susceptibility loci for *S. aureus* infection recorded in the GWAS catalog to GWAS of UKB cases defined from SGSS linkage. RSID: reference SNP cluster ID; CHR: chromosome; POS: position; OR: odds ratio.

**Supplementary Table S5:** Numbers of cases of *S. pneumoniae* cases identified by SGSS linkage (all specimen types), cross-classified by their co-identification by HES linkage, and of those, diagnosed as septicaemia by ICD-10 code.

|  | Cases defined by SGSS linkage | Cases co-identified by HES linkage | Septicaemia cases by HES linkage |
| --- | --- | --- | --- |
| All specimens defined by SGSS linkage | 1928 | 239 | 51 |
| Blood specimens defined by SGSS linkage | 222 | 128 | 48 |

**Supplementary Table S6:**
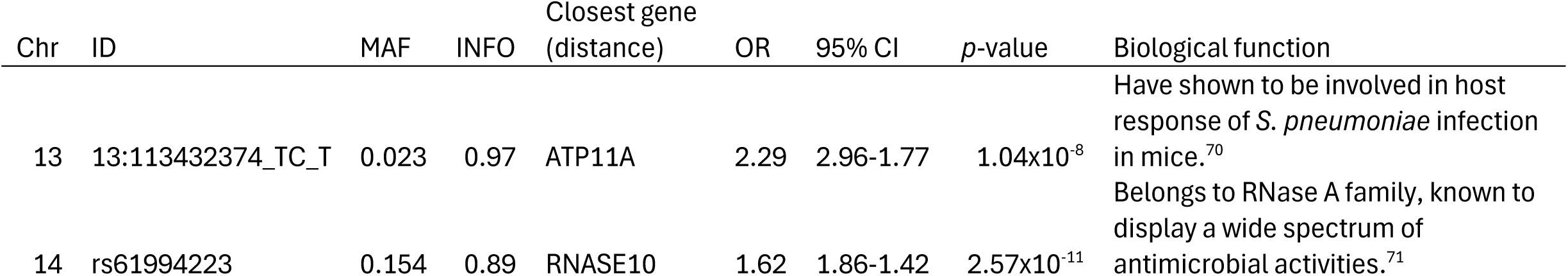
Genome-wide significant susceptibility loci for *S. pneumoniae* infection, identified by HES linkage. Chr: chromosome; RSID: reference SNP cluster ID; MAF: minor allele frequency; INFO: imputation information score; OR: odds ratio; CI: confidence interval.

**Supplementary Table S7:**
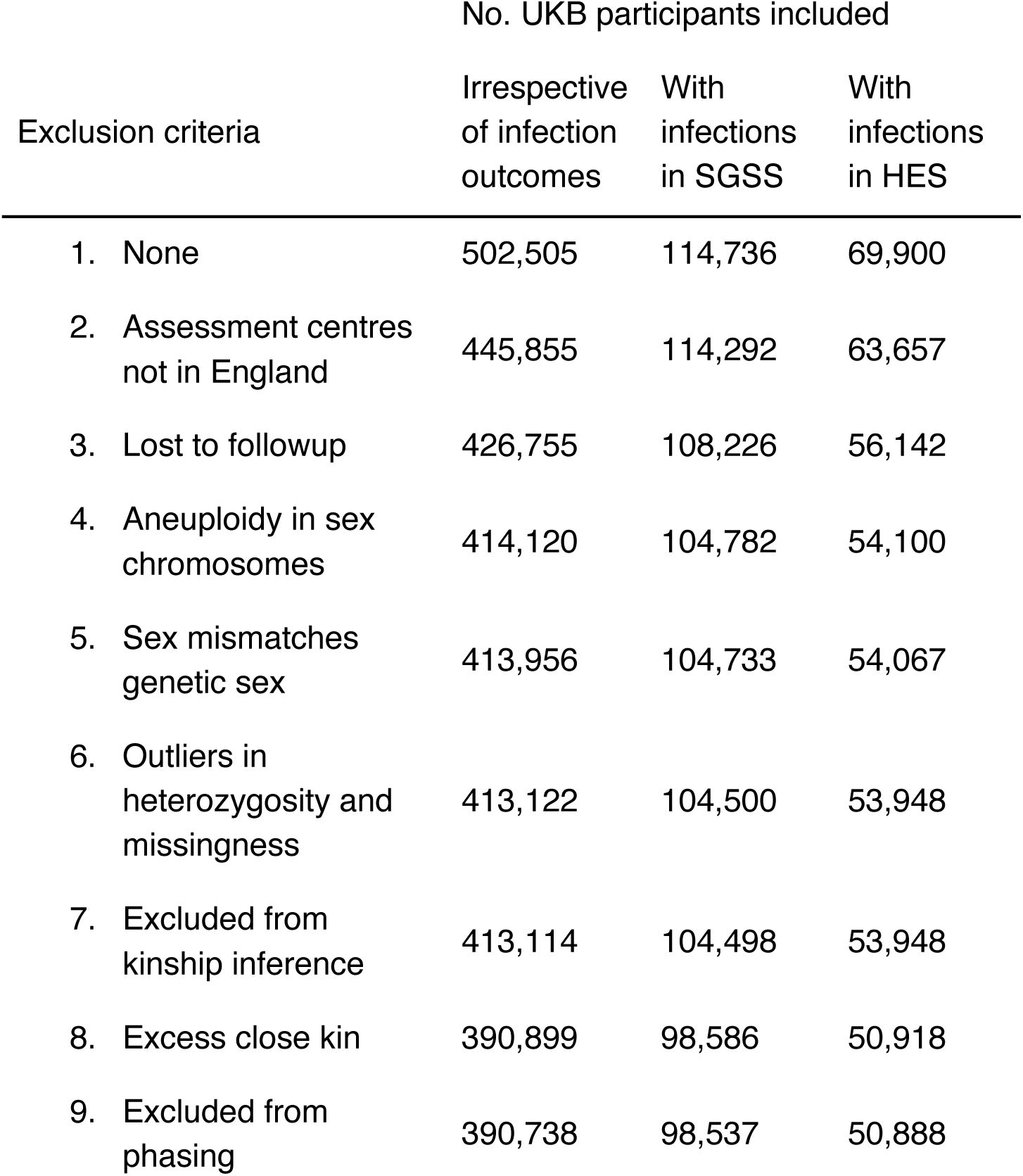
Effect of the exclusion criteria on the number of UKB participants included in the analysis.

| Exclusion criteria | No. UKB participants included |  |  |
| --- | --- | --- | --- |
|  | Irrespective of infection outcomes | With infections in SGSS | With infections in HES |
| 1. None | 502,505 | 114,736 | 69,900 |
| 2. Assessment centres not in England | 445,855 | 114,292 | 63,657 |
| 3. Lost to followup | 426,755 | 108,226 | 56,142 |
| 4. Aneuploidy in sex chromosomes | 414,120 | 104,782 | 54,100 |
| 5. Sex mismatches genetic sex | 413,956 | 104,733 | 54,067 |
| 6. Outliers in heterozygosity and missingness | 413,122 | 104,500 | 53,948 |
| 7. Excluded from kinship inference | 413,114 | 104,498 | 53,948 |
| 8. Excess close kin | 390,899 | 98,586 | 50,918 |
| 9. Excluded from phasing | 390,738 | 98,537 | 50,888 |

